# Automated analysis of lexical features in Frontotemporal Degeneration

**DOI:** 10.1101/2020.09.10.20192054

**Authors:** Sunghye Cho, Naomi Nevler, Sharon Ash, Sanjana Shellikeri, David J. Irwin, Lauren Massimo, Katya Rascovsky, Christopher Olm, Murray Grossman, Mark Liberman

## Abstract

We implemented an automated analysis of lexical aspects of semi-structured speech produced by healthy elderly controls (n=37) and three patient groups with frontotemporal degeneration (FTD): behavioral variant FTD (n=74), semantic variant primary progressive aphasia (svPPA, n=42), and nonfluent/agrammatic PPA (naPPA, n=22). Based on previous findings, we hypothesized that the three patient groups and controls would differ in the counts of part-of-speech (POS) categories and several lexical measures. With a natural language processing program, we automatically tagged POS categories of all words produced during a picture description task. We further counted the number of *wh*-words, and we rated nouns for abstractness, ambiguity, frequency, familiarity, and age of acquisition. We also computed the cross-entropy estimation, which is a measure of word predictability, and lexical diversity for each description. We validated a subset of the POS data that were automatically tagged with the Google Universal POS scheme using gold-standard POS data tagged by a linguist, and we found that the POS categories from our automated methods were more than 90% accurate. For svPPA patients, we found fewer unique nouns than in naPPA and more pronouns and *wh*-words than in the other groups. We also found high abstractness, ambiguity, frequency, and familiarity for nouns and the lowest cross-entropy estimation among all groups. These measures were associated with cortical thinning in the left temporal lobe. In naPPA patients, we found increased speech errors and partial words compared to controls, and these impairments were associated with cortical thinning in the left middle frontal gyrus. bvFTD patients’ adjective production was decreased compared to controls and was correlated with their apathy scores. Their adjective production was associated with cortical thinning in the dorsolateral frontal and orbitofrontal gyri. Our results demonstrate distinct language profiles in subgroups of FTD patients and validate our automated method of analyzing FTD patients’ speech.

## 1. Introduction

Speech production is a complex, intentional, planned activity. Speakers select appropriate words from their lexicon that are consistent with the meaning of an intended message, arrange words in a specific order following the syntactic rules of the language, plan their articulations, and articulate the prepared message following the phonological rules of the language. This involves multiple brain regions, and we can expect patients with degenerative brain conditions to show impaired speech compared to healthy adults. Moreover, depending on the form of disease, we can expect distinct impairment profiles. In this study, we investigate linguistic impairments in patients with frontotemporal degeneration (FTD) by implementing a fully automated method of lexical analysis.

FTD refers to a group of disorders caused by atrophy in the brain’s frontal, temporal, and parietal lobes, which is related to the underlying accumulation of abnormal Tau or TDP proteins. The disorders we investigated include two forms of primary progressive aphasia (PPA), the semantic variant PPA (svPPA) and the nonfluent/agrammatic variant PPA (naPPA). We also examined behavioral variant frontotemporal dementia (bvFTD). Patients with svPPA, also known as semantic dementia, are characterized by semantic impairment and difficulties in confrontation naming and lexical retrieval (Amici et al., 2007; Hodges & Patterson, 2007). Previous studies have shown that svPPA patients have difficulty processing words denoting concrete objects (Bonner, Price, Peelle, & Grossman, 2016; Bonner et al., 2009; Breedin, Saffran, & Coslett, 1994; Cousins, York, Bauer, & Grossman, 2016; Cousins, Ash, Irwin, & Grossman, 2017; Macoir, 2009), but their prosody and syntax are less disrupted (Adlam, Bozeat, Arnold, Watson, & Hodges, 2006, Ash et al. 2006; Ash et al., 2009; Nevler, Ash, Irwin, Liberman, & Grossman, 2019; Thompson & Mack, 2014). It has also been observed that svPPA patients’ lexical retrieval is related to word familiarity and frequency (Bird et al., 2000; Hodges & Patterson, 2007; Rogers, Patterson, Jefferies, & Lambon Ralph, 2015). Patients with naPPA, also known as progressive non-fluent aphasia, present with effortful speech, slow speech rate, grammatical simplification, and speech errors or apraxia of speech (AoS) (Ash et al., 2009; Grossman, 2012; Grossman et al., 1996; Josephs et al., 2006; Ogar, Dronkers, Brambati, Miller, & Gorno-Tempini, 2007). These patients may also have difficulty retrieving verbs (Hillis, Oh, & Ken, 2004; Hillis, Tuffiash, & Caramazza, 2002; Rhee, Antiquena, & Grossman, 2001). Patients with bvFTD undergo changes in personality and social cognition and also present impairments in behavior, such as apathy and disinhibition. Previous studies have reported that bvFTD patients have subtle linguistic deficits with reduced retrieval of abstract words, reduced speech rate, tangential speech with irrelevant subject matter, and limited narrative expression (Ash et al., 2006; Cousins et al., 2017; Farag et al., 2010; Gunawardena et al., 2010; Hardy et al., 2016).

While valuable, most previous studies have relied on subjective, manual assessments of speech, which require a substantial amount of time, labor, and cost. There are also potential difficulties with manually coding the part of speech (POS) categories of every token due to the time, effort, and expertise that are required, so previous studies involving POS analysis have rarely examined every word of an utterance. This is a problem in studying language use in patients with dementia, because many previous studies have shown that such patients tend to produce fewer words than controls (e.g., Ash et al. 2013; Slegers et al., 2018; Tappen et al., 2002). However, previous studies have failed to show in detail which POS categories were reduced in which patient groups due to the effort required for manual POS tagging. As a result, large-scale studies have rarely been performed. The present study describes implementation of a novel, quantitative, reproducible, automated approach to studying lexical characteristics of patients with FTD. We show that our novel methods are reliable with validation against manual gold-standard data. We also provide novel findings by directly examining all POS categories from a semi-structured speech sample elicited during a picture description task. Few studies have compared FTD subgroups on a variety of lexical measures and studied POS production in bvFTD; this is the first comprehensive assessment of POS expression in bvFTD of which we aware. We further focus on lexical characteristics of FTD patients’ speech because the lexicon is important in verbal communication where the goal is to convey meaningful messages to interlocutors. We also examine two global text measures: cross-entropy and lexical diversity. Cross-entropy is a useful measure in understanding how predictable a text sample is, in comparison to much larger language samples, and lexical diversity represents the diversity in a speaker’s vocabulary usage. Our novel, automated technique for text analysis is based on a modern natural language processing (NLP) program and examines speech samples in a large cohort of FTD patients.

Based on previous findings, we hypothesize the following

- Frequencies of POS categories as determined by an automated POS tagger and lexical measures are valuable in distinguishing the svPPA, naPPA, and bvFTD patient groups.
  - In svPPA, we expect that patients would produce fewer nouns but more pronouns than the other patients related to their impairment in confrontation naming. We also expect these patients to produce more *wh*-words (e.g., “What is this?”), since they have difficulty retrieving the names of objects or understanding a pictured object. We also expect that their nouns would be different on some lexical measures from those produced by the other patient groups due to their semantic impairment. Also, because their speech includes more pronouns and abstract, ambiguous nouns, we expect the cross-entropy measure to be low, indicating more predictability. Furthermore, we expect these language characteristics to be related to regions of cortical thinning in the temporal lobe.
  - We expect that naPPA patients would differ from the other patient groups in their frequency of speech errors, partial words, due to AoS and their difficulty in retrieving verbs. We also expect these measures to be related to cortical thinning.
  - In bvFTD, we expect to find reduced production of abstract words compared to the other groups. We also expect that bvFTD patients who are apathetic would not modify or elaborate on the details of objects, so bvFTD patients’ use of fewer adjectives was expected to be related to level of apathy. Adverb counts might also be lower in apathetic bvFTD patients, but to a lesser degree than adjective counts, since adverbs do not always serve the same modifying and elaborating role that adjectives do. Also, we expect these measures will be related to cortical thinning in the frontal lobe.
  - We expect all patients to differ from controls in lexical diversity, consistent with previous studies, which have often showed significantly decreased lexical diversity in brain-damaged patients compared to controls (e.g., Kavé and Dassa, 2018).

## 2. Methods

### 2.1 Participants

We examined 138 patients with FTD diagnosed by experienced neurologists (M.G., D.J.I.) in the Department of Neurology at the Hospital of the University of Pennsylvania according to published criteria (Gorno-Tempini et al., 2011; Rascovsky et al., 2011). This includes 42 patients with svPPA, 22 patients with naPPA, and 74 patients with bvFTD. Among the svPPA patients, we included 32 cases with concomitant mild behavioral features, a common co-occurrence. These patients did not differ significantly from the other 10 svPPA patients without behavioral impairment in terms of demographic characteristics or linguistic performance. We also included 37 healthy seniors as a control group. The Institutional Review Board of the Hospital of the University of Pennsylvania approved the study, and written consent was obtained from all participants.

All participants (n=175) were native speakers of English. The participants were matched on education level, but not on age and sex ratio (Table 1). A Tukey’s post-hoc test of the ANOVA analysis revealed that bvFTD patients were significantly younger than naPPA patients and controls (vs. naPPA, *p*=0.002; vs. control, *p*=0.007). svPPA patients were also significantly younger (vs. naPPA, *p*=0.007; vs. control, *p*=0.029). Separate chi-squared tests indicated that there were more females in the control group than in the bvFTD group (*p*=0.006) although the sex ratio was not different among the patient groups. Patient groups were matched on disease duration (*p*=0.24) and Mini Mental State Exam (MMSE, *p*=0.47).

**Table 1:** Group means (SD) and omnibus test results of clinical and demographic characteristics. ANOVA analyses were used to compare all measures between groups except sex ratio, where a chi-squared test was used. MRI: Magnetic resonance imaging, BNT: Boston Naming Test, PPT: Pyramids and Palm Trees Test, PBAC: The Philadelphia Brief Assessment of Cognition (0=most apathetic, 4=least apathetic). Numbers in square brackets are Ns when less than the total.

|  | control<br>(N=37) | bvFTD<br>(N=74) | naPPA<br>(N=22) | svPPA<br>(N=42) | Group<br>comparisons |
| --- | --- | --- | --- | --- | --- |
| **Sex** Female (N,<br>percent) | 24<br>(64.9%) | 26 (35.1%) | 11 (50%) | 23 (54.8%) | $\chi^2=9.9$,<br>$p=0.019$ |
| Male (N, percent) | 13<br>(35.1%) | 48 (64.9%) | 11 (50%) | 19 (45.2%) |  |
| **Education** | 15.9 (2.5) | 15.8 (2.8) | 15.3<br>(3.1) | 15.1 (2.8) | $F(3,171)=0.9$,<br>$p=0.437$ |
| **Age (years)** | 68.5 (7.9) | 63.1 (8.7) | 70.4<br>(9.4) | 63.3 (7) | $F(3,171)=7.3$,<br>$p<0.001$ |
| **Disease duration (years)** | - | 4.4 (3.5) | 3.2 (1.9) | 3.9 (2) | $F(2,135)=1.5$,<br>$p=0.239$ |
| **Time between MRI &**<br>**picture description**<br>**recording (months)** | - | [42] | [8] | [26] | $F(2,73)=1.1$,<br>$p=0.326$ |
|  | - | 2.2 (1.9) | 1.7 (1.7) | 2.8 (2.6) |  |
| **Mini mental state exam**<br>**(0-30)** | [31] | [68] | [20] | [38] | $F(3,153)=12.1$,<br>$p<0.001$ |
|  | 29.2 (1) | 23.6 (5.5) | 22.7 (6) | 22.1 (6.3) |  |
| **BNT (0-30)** | [23] | [68] | [16] | [40] | $F(3,143)=99.8$,<br>$p<0.001$ |
|  | 27.9 (2.5) | 23.8 (5.8) | 24.7<br>(4.6) | 7.5 (6.4) |  |
| **Animals and Tools (Max**<br>**60 sec)** | [23] | [65] | [16] | [39] | $F(3,139)=30.8$,<br>$p<0.001$ |
|  | 16.8 (4.6) | 9.2 (5.2) | 8.2 (4.4) | 5.1 (3.8) |  |
| **PPT (0-52)** | [18] | [35] | [7] | [19] | $F(3,75)=11.4$,<br>$p<0.001$ |
|  | 50.8 (1.9) | 42.9 (7.9) | 48.4<br>(2.9) | 39.6 (6.6) |  |
| **PBAC Apathy (0-4): N** | [6] | [62] | [14] | [37] | $F(3,115)=2.88$,<br>$p=0.039$ |
|  | 3.3 (0.5) | 2.1 (1.1) | 2.7 (1.2) | 2.5 (1.2) |  |

**Table 2:** Group means (SD) and omnibus test results from ANCOVA analyses of the POS categories per 100 words, total number of words, and the ratio of content words of all participants.

|  |  | Control | bvFTD | naPPA | svPPA | F | *p* |
| --- | --- | --- | --- | --- | --- | --- | --- |
| Significant group differences | Unique nouns | 14.7<br>(3.19) | 14.87<br>(5.93) | 16.73<br>(5.96) | 12.21<br>(5.19) | F(3,169)<br>=3.46 | 0.018 |
|  | Nouns | 20.32<br>(4.4) | 20.16<br>(6.48) | 21.92<br>(8.7) | 17.49<br>(5.3) | F(3,169)<br>=2.52 | 0.058 |
|  | Pronouns | 7.33<br>(2.41) | 7.13<br>(3.77) | 6.46<br>(3.2) | 9.74<br>(3.9) | F(3,169)<br>=7.66 | <0.001 |
|  | *wh*-words | 0.34<br>(0.53) | 0.6<br>(1.12) | 0.34<br>(0.99) | 1.61<br>(1.72) | F(3,169)<br>=9.26 | <0.001 |
|  | Tense-inflected<br>verbs | 12.47<br>(1.83) | 12.94<br>(3.68) | 11.26<br>(3.2) | 14.14<br>(2.98) | F(3,169)<br>=3.92 | 0.01 |
|  | Verbs | 22.56<br>(3.42) | 23.59<br>(4.86) | 20.22<br>(4.42) | 24.44<br>(4.06) | F(3,169)<br>=3.86 | 0.011 |
|  | Speech<br>errors/partial<br>words | 0.48<br>(0.89) | 1.42<br>(2.26) | 3.67<br>(3.4) | 0.89<br>(1.54) | F(3,169)<br>=4.18 | 0.007 |
|  | Adverbs | 5.59<br>(2.07) | 6.04<br>(4.36) | 4.37<br>(3.61) | 7.05<br>(3.36) | F(3,169)<br>=2.82 | 0.041 |
|  | Total words | 174.38<br>(66.38) | 109.99<br>(62.35) | 91<br>(55.8) | 127.57<br>(66.5) | F(3,169)<br>=11.37 | <0.001 |
|  | Adjectives | 5.54<br>(1.82) | 3.98<br>(3.16) | 3.17<br>(2.03) | 3.69<br>(2.04) | F(3,169)<br>=5.87 | <0.001 |
|  | Prepositions | 9.96<br>(1.94) | 7.63<br>(4.06) | 5.98<br>(3.19) | 7.24<br>(3.72) | F(3,169)<br>=7.66 | <0.001 |
| No group<br>differences | Determiners | 14.16<br>(2.48) | 14.85<br>(4.33) | 14.34<br>(5.4) | 13.35<br>(4.98) | F(3,169)<br>=0.97 | 0.41 |
|  | Conjunctions | 4.43<br>(1.91) | 5.12<br>(2.69) | 5.9<br>(4.68) | 4.85<br>(2.88) | F(3,169)<br>=1.41 | 0.24 |
|  | Fillers | 5.5<br>(2.56) | 5.89<br>(3.9) | 10.03<br>(10.3) | 6.27<br>(4.83) | F(3,169)<br>=1.46 | 0.23 |
|  | Ratio of<br>content to<br>function<br>words | 1.32<br>(0.22) | 1.36<br>(0.33) | 1.3 (0.6) | 1.32<br>(0.36) | F(3,169)<br>=0.7 | 0.55 |

We also measured patients’ performance on neuropsychological assessments (Table 1) with the Boston Naming Test (BNT, Kaplan, Goodglass, & Weintraub, 2001), Pyramids and Palm Trees Test (PPT, Howard & Patterson, 1992), Animals and Tools Category Naming Fluency (Lezak, Howieson, & Loring, 1983) to assess semantic knowledge, and the Philadelphia Brief Assessment of Cognition (PBAC, Libon et al., 2011) to assess the degree of apathy in participants. As expected, on the BNT, in which participants are asked to name an object, svPPA patients had significantly lower scores than the other groups (*p*<0.001 for all three pairwise comparisons). Patients with bvFTD also scored significantly lower on the BNT than healthy controls (*p*=0.01). On PPT, where participants were asked to choose one of two words that was more closely related in meaning to a target word, svPPA patients had lower scores than controls (*p*<0.001) and naPPA patients (*p*=0.012), and bvFTD patients also scored lower than controls (*p*<0.001). All patient groups performed poorly on the category fluency tasks, where participants were asked to name items in a given category (either animals or tools), compared to controls (*p*<0.001 for all three pairwise comparisons). The difference in the fluency task scores between bvFTD and svPPA patients was also significant (*p*<0.001). On the PBAC apathy scale, where the degree of apathy is assessed by interviewing family members or observing patients’ behavior during the clinical interview (0=most apathetic, 4=least apathetic), the result of an ANOVA analysis was significant (F(3,115)=2.88, *p*=0.039). While pair-wise group comparisons were not significant, bvFTD patients were numerically the most apathetic group. The participants’ demographic and neuropsychological characteristics are summarized in Table 1.

### 2.2 Picture description procedure

The participants were asked to describe the Cookie Theft picture from the Boston Diagnostic Aphasia Examination (Goodglass & Kaplan, 1983), and the descriptions were digitally recorded. Patients were prompted to continue describing the picture, if necessary, following a silence of several seconds, and they were encouraged to continue up to about 60 seconds after the beginning of the description. Recordings were orthographically transcribed by a linguist (S.A.), blinded to the clinical features and group membership of the participants, and further reformatted and time-stamped by trained, blinded annotators at the Linguistic Data Consortium (LDC) of the University of Pennsylvania. We note that no part of the study procedures or analyses were pre-registered prior to the research being conducted.

### 2.3 POS tagging

We employed spaCy (Honnibal & Johnson, 2015; https://spacy.io), an NLP library in Python, to automate the POS tagging process. spaCy has two different schemes of POS tagging. One is the OntoNotes 5 (Weischedel et al. 2013) version of the Penn Treebank tag set (Marcus, Santorini, & Marcinkiewicz, 1993). The other is the Google Universal POS tag set (Petrov, Das, & McDonald, 2012), which is simpler than the Penn Treebank scheme. The two POS tag schemes are not independent of each other, since spaCy maps the Penn Treebank tag to the simpler Google Universal POS tag set. Here we report the Universal POS tag results except for the calculation of the number of tense-inflected verbs, for which we used the Penn Treebank tags, because tense-inflected verbs are not distinguished by the broader Universal POS categories. The POS lists are included in the Appendix (Table A).

We wrote a Python program (S.C.) by which spaCy automatically tokenized each utterance in the transcripts with its default language model and annotated the POS category and the lemma for each word. In total, we had 21,990 tokenized words with both the Universal and Penn Treebank tags. The token count of each POS category (both Universal and Penn Treebank schemes) was tallied for each participant, and the number of each POS category per 100 words was calculated. We used POS counts per 100 words in all statistical analyses.

The Universal POS annotation scheme of spaCy uses “X” to tag words that do not exist in its language model. For example, *sptrkljgl* would be tagged as X, since the token is not a valid English word. Patients did not produce many non-English words during the picture description task, but they produced many partial words and speech errors, which looked like non-English words in the transcription. For example, in the utterance, “There’s a ***pu-***um a plate,” ***pu****-*was tagged as X by spaCy, since this is not an English word. We compared the frequency of this category by group in order to evaluate the frequency of speech errors and partial words in naPPA patients compared with other groups.

We also calculated the number of tense-inflected verbs per 100 words, the number of unique nouns per 100 words, the number of *wh*-words per 100 words and the total number of words in each speech sample, using the Penn Treebank POS tags and lemma counts. First, we summed all tokens produced by each participant for the total number of words. This measure included partial words and speech errors. The number of tense-inflected verbs was calculated by summing the number of modal auxiliary verbs, the number of past tense verbs, and the number of present tense verbs, using the Penn Treebank POS tags (Appendix Table A). This sum was used to compute the number of tense-inflected verbs per 100 words. We counted the number of unique lemmas in each speech sample and calculated the number of unique nouns per 100 words. We also counted the number of *wh*-words, “what” and “who”, using a Python script, and calculated the number of *wh*-words per 100 words to examine the clinical observations that svPPA patients use more *wh*-words to ask objects’ names than the other groups do, due to their impairments in object knowledge. To see if the ratio of POS categories differed by group, we calculated the ratio of content words to function words for each participant. The calculated measures were used for between-group comparisons, covarying for age and sex.

### 2.4 Lexical measures

We performed additional analyses of nouns because of their potential value in distinguishing FTD patient groups. We rated nouns for abstractness on a continuum from concrete to abstract (Brysbaert, Warriner, & Kuperman, 2014), semantic ambiguity (number of a given word’s meanings in a context, Hoffman, Lambon Ralph, & Rogers, 2013), word frequency (defined as word frequency per million words on a log_10_ scale, Brysbaert & New, 2009), age of acquisition (Brysbaert, Mandera, & Keuleers, 2018) and word familiarity (z-standardized measure of the number of people who know a given word, Brysbaert et al., 2018). We wrote a Python program (S.C.) to provide these parameters automatically for all nouns that spaCy annotated. We built a pipeline in the program which (1) rated a word if it was listed in the published database and (2) rated the lemma of a word if the word was not listed in the published database but its lemma was (e.g., overflowed ⇒ overflow). The program excluded a word if neither the word nor its lemma was included in the lists (e.g., *countertop, Mary Jane*). This excluded about 3% of the words tagged as nouns (141 out of 4,157 words) from the analysis. The abstractness ratings ranged from 1 to 5, where the most concrete was 5 and the most abstract was 1. For clearer representation, we inverted the scale so that the most concrete was 1 and the most abstract was 5.

Along with these measures, we also computed cross-entropy estimation using all the words of the participants’ speech. Cross-entropy estimation is a measurement that estimates the predictability of all words of a document with respect to their predictability in a larger language sample. For example, high cross-entropy (uncertainty) is observed in a document that uses unusual words given the source language sample. A computational linguist (M.L.) computed the cross-entropy estimation of the speech samples by patients, based on a 1-gram language model of three large-scale corpora: the SUBTLEXus (Brysbaert & New, 2009), Fisher English Training Speech (Cieri, Graff, Kimball, Miller, & Walker, 2004), and Switchboard (Godfrey & Holliman, 1997).

We also calculated lexical diversity for each patient. Traditionally, lexical diversity has been measured using the type/token ratio, where *type* is the number of unique words and *token* is the number of instances of each word. However, the type/token ratio has the disadvantage that the measure is affected by the total number of words. To address this problem, various approaches have been suggested by previous studies (e.g., Covington & McFall, 2010; Jarvis, 2002; McKee, Malvern, & Richards, 2000; Moscoso del Prado Martín, 2017; Tweedie & Baayen, 1998). In this study, we used the moving-average type/token ratio (Covington & McFall, 2010), which has been reported to be a stable measure for lexical diversity (Cunningham & Haley, 2020). It calculates a type/token ratio for a fixed-length window, moving one word at a time from the beginning to the end of a text document, and averages type/token ratios from all windows. We varied the length of the window from 20 to 35 words by 5-word increments. Since the results were the same regardless of the window size, we reported results from 20-word windows in Figure 2 and Table 3. Hereafter, abstractness, ambiguity, frequency, familiarity, AoA, cross-entropy, and lexical diversity are referred to as “lexical measures”. “Language measures” is used to refer to both POS counts and the lexical measures.

**Table 3:** Group means (SD) and omnibus test results from ANCOVA analyses of the lexical measures. AoA: Age of acquisition.

|  | Control | bvFTD | naPPA | svPPA | F | *p* |
| --- | --- | --- | --- | --- | --- | --- |
| Abstractness (noun) | 1.52 (0.76) | 1.55 (0.83) | 1.4 (0.59) | 1.92 (1.14) | F(3,169)=11.68 | <0.001 |
| Ambiguity (noun) | 1.65 (0.25) | 1.64 (0.26) | 1.64 (0.23) | 1.74 (0.28) | F(3,169)=11.01 | <0.001 |
| Frequency (noun) | 3.39 (0.86) | 3.52 (0.91) | 3.44 (0.91) | 3.94 (0.95) | F(3,169)=12.99 | <0.001 |
| Familiarity (noun) | 2.38 (0.14) | 2.38 (0.16) | 2.39 (0.14) | 2.4 (0.16) | F(3,169)=3.81 | 0.011 |
| AoA (noun) | 4.51 (1.42) | 4.36 (1.33) | 4.21 (1.24) | 4.15 (1.13) | F(3,169)=4.27 | 0.005 |
| Cross-entropy | 9.72<br>(0.49) | 9.61<br>(0.66) | 9.9 (0.84) | 9.1 (0.79) | F(3,169)=7.7 | <0.001 |
| Lexical<br>diversity | 0.85<br>(0.03) | 0.79<br>(0.09) | 0.79<br>(0.06) | 0.81 (0.09) | F(3,169)=6.21 | <0.001 |

**Figure 1:**
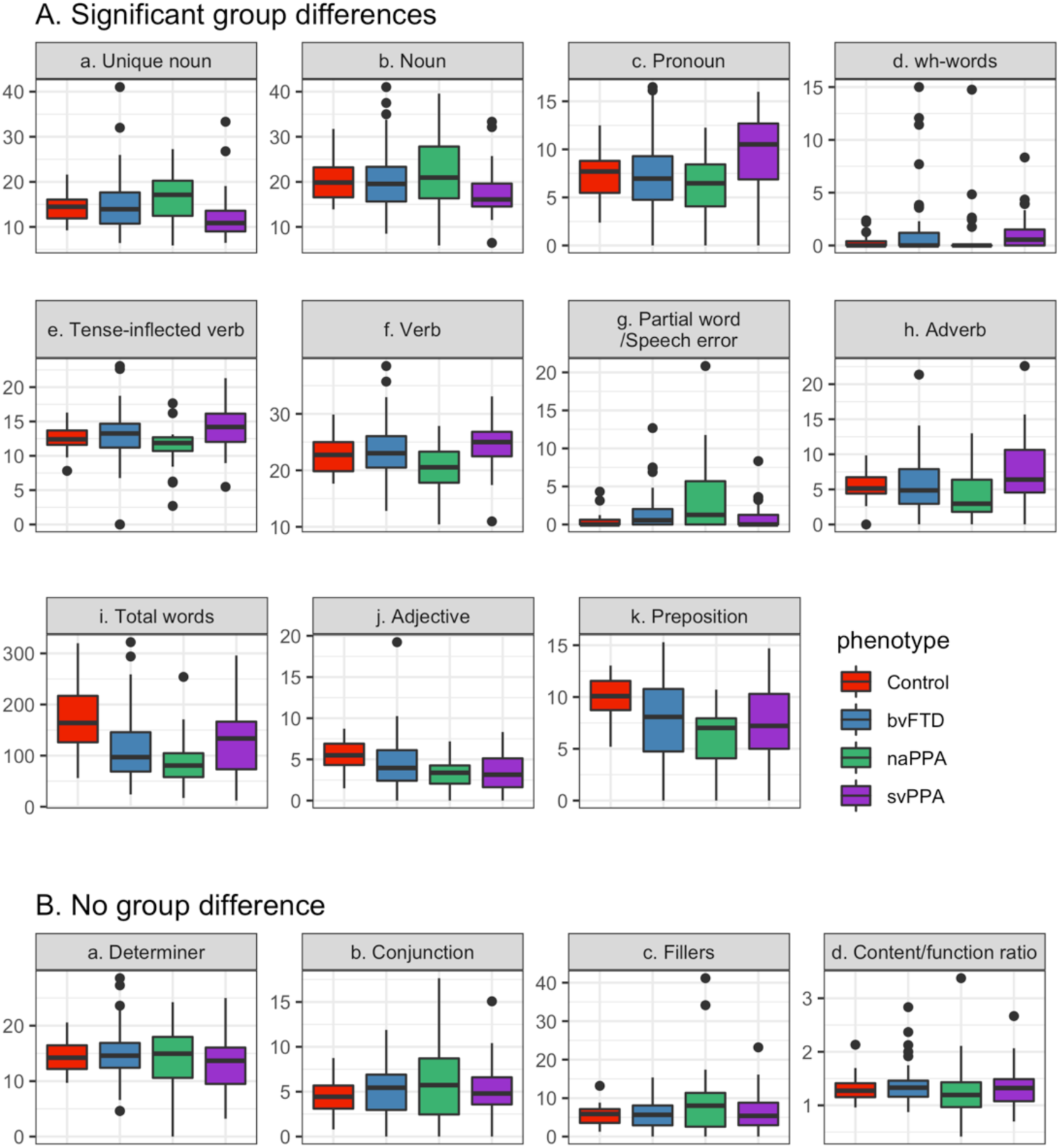
Median, 1SD, 25th-75th percentile and outliers in POS categories per 100 words, total number of words and the ratio of content words by phenotype.

**Figure 2:**
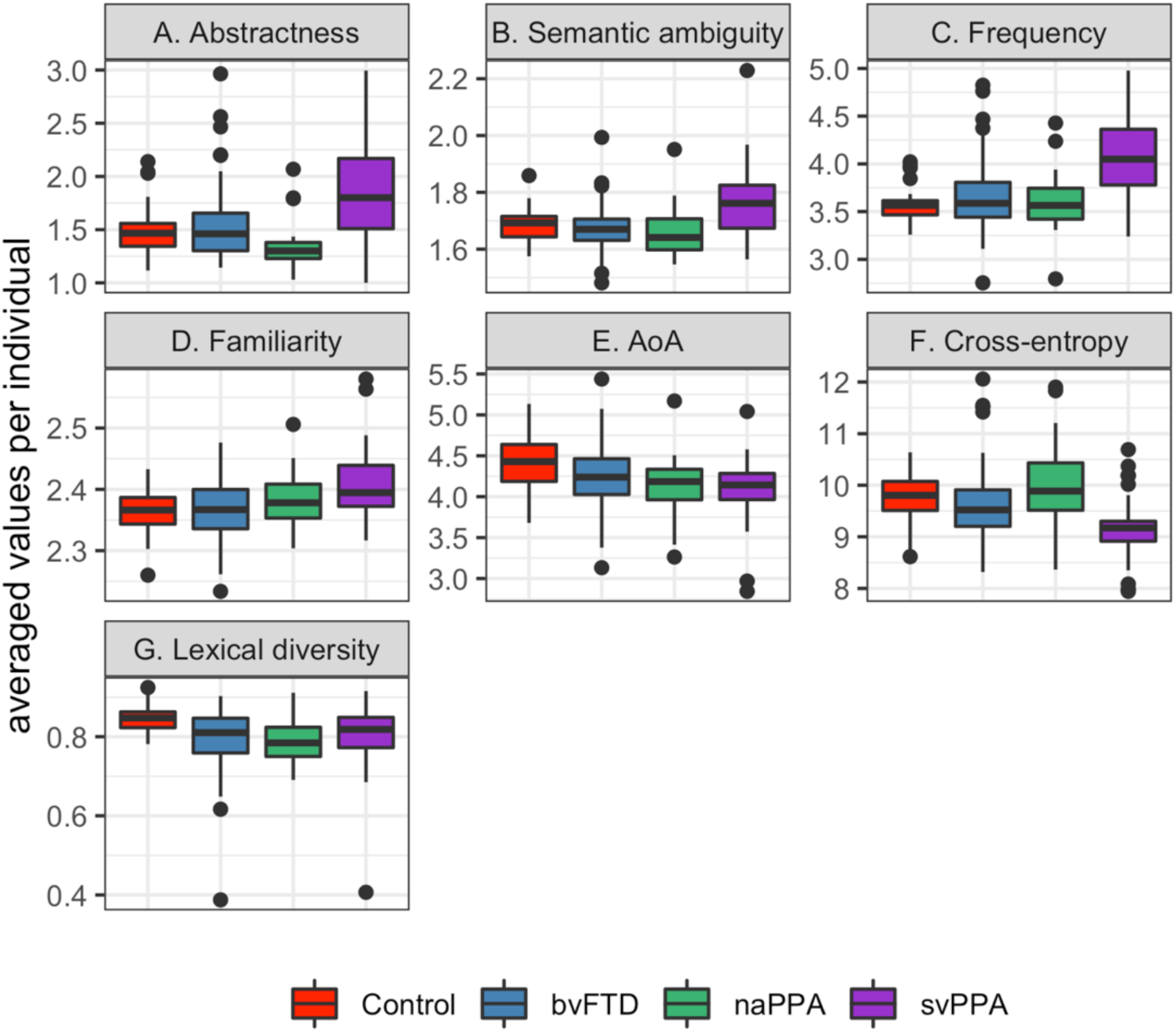
Median, 1SD, 25th-75th percentile and outliers of abstractness scores, semantic ambiguity ratings, word frequency, word familiarity, and age of acquisition of nouns; and cross-entropy estimation and lexical diversity across all words.

### 2.5 Imaging methods

High resolution T1 volumetric brain MRI data that were collected on a Siemens 3.0T Trio scanner at 1mm isotropic resolution were available for a subset of patients (n=94): 18 controls, 42 bvFTD, 8 naPPA, and 26 svPPA patients. The mean time interval between MRI and speech sample collection was 1.95 months (SD=2.11 months). Clinical and demographic characteristics of this subset of patients matched those of the patients in the full dataset, and the groups in this subset were matched on demographic characteristics. The demographic and language measurements of these patient groups are summarized in the Appendix (Tables B-C).

Sixty-five images were collected in an axial plane with repetition time=1620 msec, echo time=3.87 msec, slice thickness=1.0 mm, flip angle=15°, matrix=192×256, and in-plane resolution=0.9766×0.9766 mm. Twenty-nine images were collected with a sagittal acquisition with repetition time=2300 msec, echo time=2.95 msec, slice thickness=1.2 mm, flip angle=9°, matrix=256×240, and in-plane resolution=1.05×1.05 mm. Briefly, whole-brain MRI volumes were preprocessed using the antsCorticalThickness.sh processing pipeline, implemented using the Advanced Normalization Tools (ANTs) (https://github.com/ANTsX/ANTs; Tustison et al., 2014). Cortical thickness was estimated at each voxel of the cortex using the DiReCT algorithm (Das, Avants, Grossman, & Gee, 2009). easy_lausanne (https://github.com/mattcieslak/easy_lausanne; Daducci et al., 2012) run on our local template, which was created based on data from the Open Access Series of Imaging Studies (OASIS) (Marcus, Fotenos, Csernansky, Morris, & Buckner, 2007), to create a standard cortical parcellation. The template parcellation was then spatially normalized to each participant’s native T1 space using the template-to-native T1 warps generated by ANTs, and then we calculated the mean cortical thickness in each region of interest (ROI) of the Lausanne250 scale, which we used for our analysis.

To identify regions of atrophy in patients, we compared cortical thickness of all patients in each patient group with those of the controls for all cortical regions of interest (ROIs) and selected our specific ROIs for each patient group, where patients’ cortical thickness was significantly thinner than that of the controls (*p*<0.01 for svPPA and bvFTD, and *p*<0.05 for naPPA, both uncorrected *p*-values). We applied a more lenient *p*-value threshold in selecting ROIs for naPPA patients due to the small number of patients with MRI data. We further identified ROIs that were significantly correlated with the degree of apathy (PBAC, Table 1) for bvFTD patients among the selected ROIs (*p*<0.05) to mask the regressions. This method enabled us to restrict our interpretation of the regression results of adjectives in bvFTD to those brain regions that were significantly related to apathy. Adverbs were not considered in the MRI analyses since the apathy scores were not significantly correlated with adverb production for bvFTD patients.

### 2.6 Statistical considerations

Since the abstractness, ambiguity, frequency, familiarity, and AoA measures were rated for each noun, we averaged those values per individual and used the mean values of the participants for group comparisons. We did not average cross-entropy and lexical diversity measures, since these were global measures (only one value per individual). Levene’s test for homogeneity of variance, residuals, and Q-Q plots were employed to validate the requirements for parametric tests. Group comparisons were performed with Analysis of Covariance (ANCOVA) for the frequency of each POS category per 100 words and each of the lexical measures as a dependent variable, with phenotype as an independent variable. We introduced age and sex as covariates in the group comparison analyses of all language measures, as the groups were not matched on these factors. For those measures where the requirements for parametric tests were not met, we performed the rank-based inverse normal transformation (Conover, 1980) on the values of language measures, and the transformed values were used as the dependent variable in an ANCOVA. When there was a significant group effect, pair-wise group comparisons were conducted with the *lsmeans* package (Lenth, 2016) in R to adjust for multiple comparisons with false discovery rate. Since the group difference from ANCOVA was marginal in the counts of nouns and adverbs per 100 words, we performed logistic regressions as supplementary analyses with age and sex as covariates to compare the number of patients who had a z-score < −1 by group, where the z-score scale was computed based on the controls’ mean and standard deviation. For the supplementary analysis for noun counts, we coded participants who produced fewer nouns (z-score < −1) as 1 and others as 0 for a dependent variable and ran a logistic regression with svPPA patients as a reference group and phenotype as an independent variable, controlling for age and sex. For the supplementary analysis of adverb counts, we coded participants who produced fewer adverbs as 1 and others as 0, with the naPPA group as our reference. We selected these reference groups based on the ANCOVA results. A Pearson’s correlation test was performed to relate adjective and adverb counts to the apathy scores on the PBAC for each group to test our hypothesis for bvFTD patients. A series of separate linear regression analyses were performed to relate the cross-entropy estimations to the five lexical measures: abstractness, ambiguity, frequency, familiarity, and AoA.

Linear regression analyses were used to relate the language measures to cortical thinning. We implemented univariate multiple regression analyses, covarying for potential confounding factors: the pulse sequence type used for MRI acquisition, patients’ age, and disease duration. We did not covary for sex because the participants with MRI data did not significantly differ in the sex ratio across groups and there was no consistent evidence of the effect of sex on cortical thinning. The regions selected for svPPA and naPPA patients were used to relate their regions of cortical thinning to language measures that significantly differed between groups. The regions that were significantly related to the apathy scores in bvFTD patients were used to relate cortical thinning to adjective counts per 100 words. We report t-statistics at a significance level of 0.05 (two-tailed, uncorrected) for these regressions. All statistical analyses were performed in R (R Core Team, 2019) version 3.5.2 and RStudio (RStudio Team, 2016) version 1.1.456 (S.C.).

## 3. Accuracy validation of spaCy POS tags

Despite the fact that the accuracy of POS tagging reported by spaCy is very high (about 97%; https://spacy.io/models/en), it was not clear how well it would perform for a clinical dataset with abnormal speech. The training data (OntoNotes 5; Weischedel et al., 2013) of spaCy included natural conversations, but the ratio of conversational speech to written texts was only around 8.3% (120K out of 1.4 million words) and the conversations were between healthy adults. To validate the accuracy of the spaCy POS tags on natural speech of a clinical population with abnormal speech, a linguist (S.A.) who was blinded to the automated analysis manually tagged a random subset of the transcripts comprehensively using the Google Universal POS scheme (6 Controls, 5 naPPA, 7 svPPA, and 7 bvFTD; 25 cases in total; 14.3% of the full dataset) to generate a gold standard dataset. We compared the results of spaCy in the same POS scheme to our gold standard dataset to calculate the error rates.

The error rate was generally low in all groups. The overall accuracy of spaCy on this subset of the picture description data was 91.1%, and the variances between the groups were not significantly different (Levene’s test for homogeneity of variance: F(3,21)=2.69, *p*=0.072). Also, a one-way ANOVA test revealed that the difference in error rates between the groups was not significant (F(3,21)=2.695, *p*=0.075). The mean error rate of the control group was 5.4% (SD=1.7%). The error rates of individual svPPA, naPPA, and bvFTD patients were slightly higher than that of the controls (svPPA: 8.8% (SD=2.8%); naPPA: 13.3% (SD=9.2%); bvFTD: 9.0% (SD=3.0%)), but the difference among the patients groups was not significant (F(2,16)=1.32, *p*=0.3). While the error rates for svPPA and bvFTD did not differ from that of controls, the difference between naPPA patients and the controls was significant (*p*=0.049). This was expected, since naPPA speech contains the largest number of speech errors and partial words (see below) and thus differs most from the training data of spaCy.

For further validation, we correlated the token counts of nouns, tense-inflected verbs, and speech errors/partial words from spaCy with the counts that a linguist (S.A.) manually coded for all 175 participants. For the correlation between the noun counts of each individual, we used all NOUN tokens in the Universal tag set. Modal auxiliaries (MD), past (VBD) and present (VBP, VBZ) tense verbs in the Penn Treebank tag set were used for the correlation with tense-inflected verb counts. For speech errors, we compared the X category in the Universal tag set with the counts of manually coded speech errors. We found that the noun and inflected verb counts of spaCy and counts of those categories in our manual coding were strongly correlated (nouns: r=0.958, *p*<0.001; verbs: r=0.973, *p*<0.001). Also, the correlation of counts of X with our manual coding of speech errors was significant (r=0.43, *p*<0.001), suggesting that the POS tags produced by spaCy were reliable.

## 4. Results

We first present the results of automatic POS tagging (Section 4.1). Next, we show the group differences in the lexical measures (Section 4.2). In Section 4.3, we present the regression results with MRI data.

### 4.1 POS categories and derived measures

Table 2 summarizes the statistical results of the POS measures per 100 words. The groups differed significantly in the number of unique nouns (Fig. 1Aa). svPPA patients produced fewer unique nouns than naPPA patients (*p*=0.022) and marginally fewer than bvFTD patients (*p*=0.056). Noun production marginally varied by phenotype after controlling for age and sex (Fig. 1Ab). However, group-wise paired comparisons failed to reach significance (svPPA vs. bvFTD: *p*=0.062; svPPA vs. naPPA: *p*=0.062). A supplementary analysis with a logistic regression revealed that there were significantly more svPPA patients who produced fewer nouns (z-score < −1) compared to bvFTD patients (*Ζ*=-2.01, *p*=0.044) and controls (*z*=-2.75, *p*=0.006) but not compared to naPPA patients (*Ζ*=-1.67, *p*=0.096). Pronoun production (Fig. 1Ac) significantly differed between groups; pronouns were more frequent for svPPA patients than for the other groups (svPPA vs. control: *p*=0.016; svPPA vs. naPPA: *p*=0.005; svPPA vs. bvFTD: *p*=0.002). Also, the groups differed in the number of *wh*-words per 100 words (Fig. 1Ad). Patients with svPPA produced more *wh*-words than the other groups (*p*<0.001 for all three pairwise comparisons).

The number of tense-inflected verbs per 100 words differed significantly by group (Fig. 1Ae). Pairwise group comparisons revealed that naPPA patients produced fewer tense-inflected verbs than svPPA patients (*p*=0.006). Similarly, the group difference in the total number of verbs was significant (Fig. 1Af). naPPA patients produced fewer verbs than svPPA patients (*p*=0.008) and bvFTD patients (*p*=0.016). The groups were also different in the counts of speech errors and partial words (Fig. 1Ag). naPPA patients produced this category significantly more frequently than controls (naPPA vs. control: *p*=0.005). Adverb production also differed by group (Fig. 1Ah). naPPA patients tended to produce fewer adverbs than svPPA patients (*p*=0.052). A supplementary analysis with logistic regression showed that the number of naPPA patients who produced fewer adverbs (z-score < −1) was greater than the number of svPPA patients (*Ζ*=-3.05, *p*=0.002) and controls (*z*=-3.57, *p*<0.001) but not greater than the number bvFTD patients (*z*=-1.8, *p*=0.07). The adverb counts per 100 words were not significantly correlated with apathy scores in any of the four groups (*p*>0.05).

The total number of words participants produced during the picture description differed significantly by group (Fig. 1Ai). Controls produced significantly more words than any of the patient groups (vs. bvFTD: *p*<0.001, vs. naPPA: *p*<0.001, vs. svPPA: *p*=0.006). Similarly, adjective production per 100 words significantly varied by group (Fig. 1Aj), and all patient groups used fewer adjectives than controls (vs. bvFTD: *p*=0.013; vs. naPPA: *p*=0.003; vs. svPPA: *p*=0.002). Furthermore, bvFTD patients’ adjective counts per 100 words were significantly correlated with their apathy scores (r=0.32, *p*=0.01). The correlations of adjective production and apathy scores were not significant in the other three groups, and bvFTD patients’ apathy scores were not significantly correlated with the other POS categories. The group difference in prepositions (Fig. 1Ak) was significant. Each patient group produced fewer prepositions than controls (vs. bvFTD: *p*=0.004; vs. naPPA: *p*<0.001; vs. svPPA: *p*=0.004). The differences among the patient groups for these categories were not significant.

The productions of conjunctions, determiners, fillers and the ratio of content to function words did not differ by group (Fig. 1B).

### 4.2 Lexical measures

All participants produced nouns that were not abstract in the picture description task, which is not surprising given the task of describing a picture that contains concrete objects. Yet, the group differences in abstractness were significant (Fig. 2A). svPPA patients produced nouns that were more abstract (i.e., less concrete) compared to bvFTD patients (*p*<0.001), naPPA patients (*p*<0.001), and controls (*p*=0.001).

Semantic ambiguity ratings of nouns also differed significantly by group (Fig. 2B). Nouns produced by svPPA patients showed higher semantic ambiguity than those produced by all other groups (vs. bvFTD: *p*<0.001; vs. naPPA: *p*<0.001, vs. controls: *p*=0.008).

Patients tended to use more frequent nouns than controls, and the group difference in the frequency of nouns was highly significant (Fig. 2C). svPPA patients produced more frequent nouns than bvFTD patients, naPPA patients, and controls (*p*<0.001 for all three pairwise comparisons).

The familiarity of nouns also significantly differed by group (Fig. 2D). svPPA patients used more familiar nouns than bvFTD patients (*p*=0.02).

All patients tended to produce nouns acquired at an earlier age than controls (Fig. 2E), and the group difference in the age of acquisition of nouns was significant. svPPA patients produced nouns that were acquired earlier than controls (*p*=0.007).

The cross-entropy estimation differed significantly by phenotype (Fig. 2F); the cross-entropy estimation of svPPA patients was lower than that of bvFTD patients (*p*=0.006), naPPA patients (*p*<0.001), and controls (*p*=0.001). In other words, words produced by svPPA patients were more predictable than those produced by the other groups. To further examine why svPPA patients’ cross-entropy estimation was lower than those of the other groups, separate linear regression analyses were performed to relate cross-entropy estimation in svPPA patients to abstractness, ambiguity, frequency, familiarity, and AoA of nouns they produced. We found that abstractness, ambiguity, and word frequency were significantly related to cross-entropy estimation in svPPA (abstractness: *β*=-0.63, *p*<0.001, word frequency: *β*=-0.88, *p*<0.001; semantic ambiguity: *β*=-2.8, *p*=0.019).

There was a significant group difference in lexical diversity that was measured by the moving-average type/token ratio with a window size of 20 words (Fig. 2G). Elderly controls showed higher lexical diversity than all patient groups (vs. bvFTD: *p<*0.001, vs. naPPA: *p*=0.019, vs. svPPA: *p=*0.019). When we tried different window sizes (25 words and 30 words), we found the same group differences (25-word window: vs. bvFTD: *p*=0.002, vs. naPPA: *p*=0.004, vs. svPPA: *p*=0.018; 30-word window: vs. bvFTD: p=0.001, vs. naPPA: p=0.006, vs. svPPA: p=0.019).

### 4.3 MRI results

Since patients showed significant differences, we examined the relations between cortical thinning and specific language measures in each group. We found distributions of cortical thinning that were representative of each group (Ash et al., 2012, 2009; Cousins et al., 2016; Massimo et al., 2009). The MRI results showed that svPPA patients had significant cortical thinning in the anterior temporal and orbital frontal cortex areas of both hemispheres, but cortical thinning was more prominent in the left hemisphere than the right hemisphere (*p*<0.01; Fig. 3A). naPPA patients had significant cortical thinning most prominently in the left middle frontal, inferior temporal and middle temporal regions, but also apparent in the left supramarginal gyrus, right temporal gyrus, and right pars opercularis (*p*<0.05, Fig. 3B). bvFTD patients had significant cortical thinning in the frontal and temporal lobes of both hemispheres (*p*<0.01; Fig. 3C). We examined patients’ speech production in relation to cortical thinning in greater detail, as summarized in Table 4. Examples of the associations are illustrated in Figure 3.

**Table 4:** Results of regression analyses with cortical thinning in patients. L: left, R: right.

| **svPPA** | **Estimate** | **Std. Error** | **t-value** | **p-value** |
| --- | --- | --- | --- | --- |
| *Noun* |  |  |  |  |
| L inferior temporal | 0.059 | 0.021 | 2.85 | 0.01 |
| L middle temporal | 0.054 | 0.026 | 2.09 | 0.049 |
| L superior temporal | 0.045 | 0.021 | 2.18 | 0.041 |
| L insula | 0.035 | 0.016 | 2.19 | 0.04 |
| *Pronoun* |  |  |  |  |
| L inferior temporal | -0.098 | 0.038 | -2.54 | 0.019 |
| L parahippocampal | -0.059 | 0.028 | -2.16 | 0.043 |
| L entorhinal | -0.104 | 0.05 | -2.08 | 0.049 |
| *Wh-words* |  |  |  |  |
| L inferior temporal | -0.219 | 0.08 | -2.6 | 0.021 |
| L middle temporal | -0.244 | 0.11 | -2.14 | 0.044 |
| L superior temporal | -0.19 | 0.087 | -2.2 | 0.039 |
| L fusiform | -0.303 | 0.108 | -2.818 | 0.01 |
| L insula | -0.142 | 0.065 | -2.207 | 0.04 |
| *Abstractness* |  |  |  |  |
| L temporal pole | -0.582 | 0.228 | -2.55 | 0.019 |
| L inferior temporal | -0.531 | 0.218 | -2.42 | 0.025 |
| L middle temporal | -0.652 | 0.225 | -2.89 | 0.011 |
| L superior temporal | -0.51 | 0.189 | -2.69 | 0.019 |
| L fusiform | -0.597 | 0.243 | -2.49 | 0.027 |
| R superior temporal | -0.309 | 0.14 | -2.21 | 0.038 |
| *Semantic ambiguity* |  |  |  |  |
| L inferior temporal | -2.609 | 0.833 | -3.11 | 0.007 |
| L middle temporal | -2.617 | 0.896 | -2.96 | 0.011 |
| L bank superior temporal | -1.795 | 0.572 | -3.13 | 0.006 |
| L superior temporal | -1.946 | 0.693 | -2.8 | 0.013 |
| L supramarginal | -1.722 | 0.601 | -2.86 | 0.018 |
| L insula | -0.5 | 0.205 | -2.39 | 0.026 |
| L lateral orbitofrontal | -1.182 | 0.564 | -2.1 | 0.048 |
| *Word frequency* |  |  |  |  |
| L inferior temporal | -0.627 | 0.258 | -2.46 | 0.024 |
| L middle temporal | -0.685 | 0.264 | -2.58 | 0.019 |
| L bank superior temporal | -0.379 | 0.176 | -2.16 | 0.043 |
| L superior temporal | -0.49 | 0.208 | -2.34 | 0.031 |
| L fusiform | -0.593 | 0.267 | -2.22 | 0.037 |
| *Word familiarity* |  |  |  |  |
| L inferior temporal | -0.755 | 0.29 | -2.61 | 0.016 |
| L middle temporal | -0.83 | 0.247 | -3.41 | 0.009 |
| L superior temporal | -0.53 | 0.182 | -2.98 | 0.018 |
| L rostral middle frontal | -0.821 | 0.216 | -3.8 | 0.001 |
| R rostral middle frontal | -0.608 | 0.222 | -2.72 | 0.014 |
| L precentral | -0.599 | 0.163 | -3.67 | 0.001 |
| L supramarginal | -0.517 | 0.19 | -2.72 | 0.013 |
| L lateral orbitofrontal | -0.365 | 0.163 | -2.24 | 0.001 |
| R superior frontal | -0.592 | 0.218 | -2.72 | 0.013 |
| R pars opercularis | -0.549 | 0.192 | -2.86 | 0.009 |
| *Cross-entropy estimation* |  |  |  |  |
| L inferior temporal | 0.451 | 0.187 | 2.4 | 0.027 |
| L middle temporal | 0.419 | 0.199 | 2.1 | 0.048 |
| L bank superior temporal | 0.348 | 0.143 | 2.45 | 0.026 |
| L superior temporal | 0.392 | 0.156 | 2.51 | 0.02 |
| L fusiform | 0.713 | 0.224 | 3.18 | 0.004 |
| **naPPA** | Estimate | Std. Error | t-value | p-value |
| *Speech errors / Partial words* |  |  |  |  |
| L rostral middle frontal | -0.194 | 0.044 | -4.39 | 0.022 |
| **bvFTD** | Estimate | Std. Error | t-value | p-value |
| *Adjectives* |  |  |  |  |
| L orbitofrontal | 0.07 | 0.028 | 2.56 | 0.015 |
| L rostral middle frontal | 0.05 | 0.024 | 2.28 | 0.031 |
| L superior frontal | 0.07 | 0.03 | 2.29 | 0.03 |
| L caudal middle frontal | 0.07 | 0.025 | 2.67 | 0.035 |
| L post central | 0.06 | 0.023 | 2.49 | 0.018 |
| R pre central | 0.07 | 0.033 | 2.22 | 0.032 |
| R post central | 0.07 | 0.025 | 2.74 | 0.009 |

**Figure 3:**
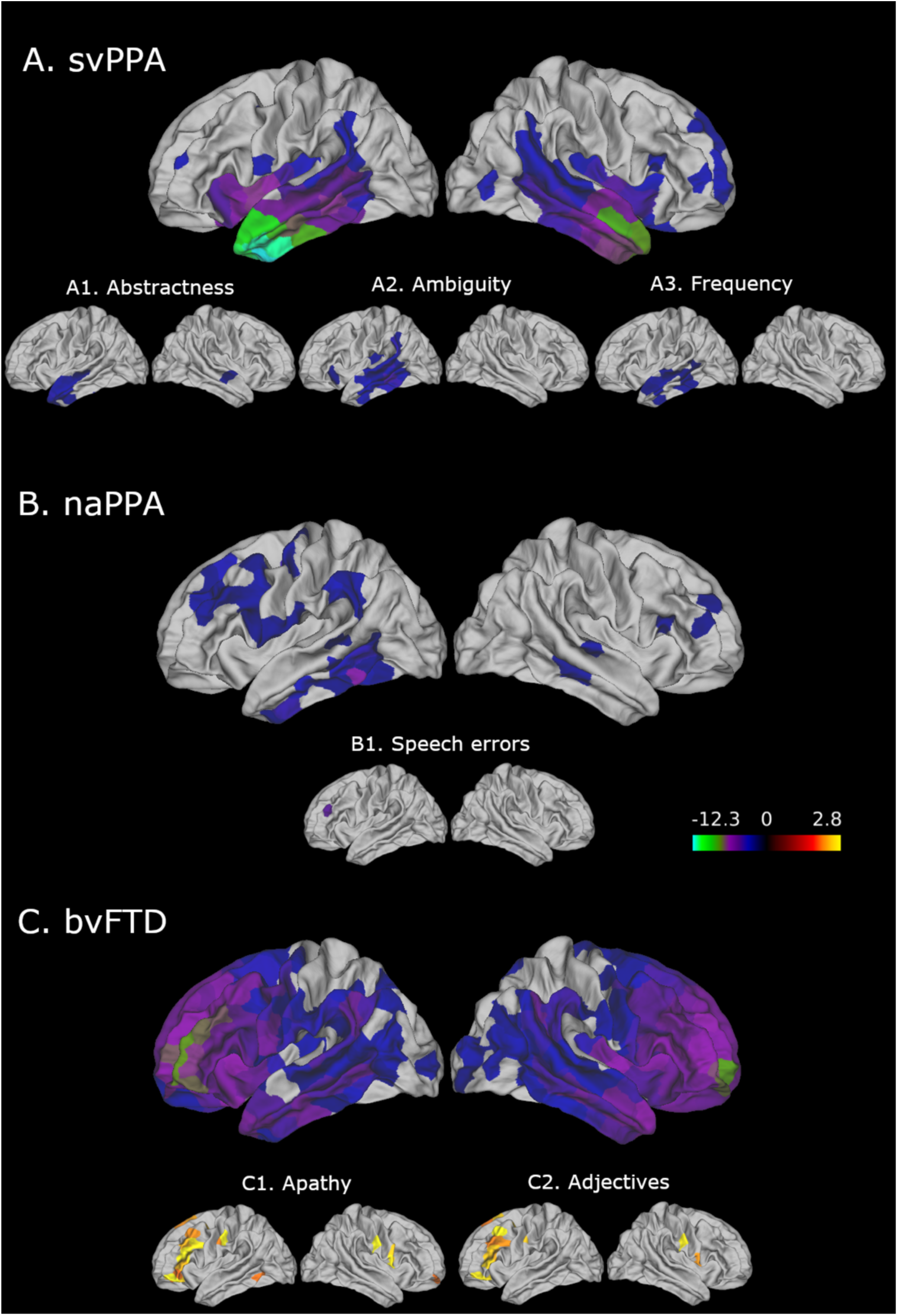
Cortical thinning in svPPA (A), naPPA (B) and bvFTD (C) patients, and areas with cortical thinning that were significantly related to linguistic measures (*p*<0.05, uncorrected) in svPPA (A1-3), naPPA (B1), and bvFTD (C2) patients. Please note that these images are for illustration, and the complete results are summarized in Table 4.

We selected the language measures that were distinctive of svPPA patients in our main analyses outlined above. These showed significant associations with cortical thinning in anterior and middle temporal regions of the left hemisphere (Table 4). Figure 3 shows brain images for the cortical thinning of abstractness, ambiguity, and frequency that are frequently described for svPPA in the literature (Fig. 3A1-3).

We also found that the production of speech errors and partial words was related to cortical thinning in the left rostral middle frontal gyrus for naPPA patients (Fig. 3B1), suggesting that speech errors and partial words are related to impairment in frontal executive functions. We also related verb, tense-inflected verb, and adverb counts to cortical thinning in naPPA patients, but the results were not significant.

The areas that showed a significant relation of cortical thinning to apathy in bvFTD patients (Fig. 3C1) are also significantly and positively related to their adjective production (Fig. 3C2). These areas include the left rostral and caudal middle frontal, the left superior frontal, and orbitofrontal regions.

## 5. Discussion

In this study, we examined word production and lexical measures of speech in FTD patients with a novel, automated method that is objective, comprehensive and reproducible. The POS counts derived from the Universal tag set were highly correlated with manually coded POS tags (Section 3). Moreover, distinct language measures were associated with each patient group (Sections 4.1–4.2). We found that svPPA patients produced fewer unique nouns than naPPA patients, and these nouns were more ambiguous, abstract, and frequent than those of naPPA and bvFTD patients. Correspondingly, svPPA patients produced more pronouns and *wh*-words. A new measure of cross-entropy estimation showed that their word selection in general was more predictable from its context than that of the other groups, and this was related in part to noun abstractness, ambiguity, and frequency. Patients’ words were less diverse than those of controls, but there was no significant group difference among the patient groups. naPPA patients produced fewer adverbs and more speech errors and partial words than the other groups. bvFTD patients produced fewer adjectives than controls, and their adjective production was significantly correlated with apathy scores. We also found significant associations between our language measures and cortical thinning. Cortical thinning in left anterior inferior and middle temporal gyri was associated with language measures in svPPA, and cortical thinning in the left middle frontal gyrus was associated with speech errors and partial words in naPPA. Cortical thinning in the left dorsolateral frontal and orbitofrontal gyri was associated with decreased adjective production in bvFTD. We discuss these findings in turn below.

### 5.1 Lexical characteristics in svPPA

The profiles of svPPA patients’ nouns exhibited characteristics that significantly differed from those of the other groups. They displayed high abstractness, semantic ambiguity, word frequency, and word familiarity. This is in line with other findings consistent with the hypothesis attributing the deficit in svPPA in part to the degradation of visual feature knowledge associated with object concepts (Bird, Lambon Ralph, Patterson, & Hodges, 2000; Bonner et al., 2016, 2009; Cousins et al., 2016; Cousins et al., 2017; Cousins, Ash, Olm, & Grossman, 2018; Hoffman et al., 2013), which is due to cortical thinning in the left anterior and inferior temporal regions of the brain. This region constitutes a portion of visual association cortex which may contribute to the representation of visual feature knowledge associated with object concepts. It may explain in part why svPPA patients produced nouns with high abstractness in our results: abstract nouns are less dependent on visual feature knowledge to derive their meaning, thereby reducing the need to activate the anterior and inferior temporal regions of the brain. We also found that an increase in the abstractness rating of nouns was related to cortical thinning in the left anterior temporal region. In the context of concrete noun difficulty due to degraded representations of visual objects, it is not surprising that svPPA patients may substitute more pronouns, and this was reflected in associations with cortical thinning in the left temporal lobe and pronoun usage.

Previous observations have showed that svPPA patients’ lexical retrieval is strongly graded by word familiarity and frequency (Bird et al., 2000; Hodges & Patterson, 2007; Rogers, Patterson, Jefferies, & Lambon Ralph, 2015). These observations suggest that at least some proportion of the svPPA patients’ picture description deficit is due in part to a lexical retrieval deficit that extends beyond their degraded semantic representations of object knowledge. As for semantic ambiguity, Hoffman et al. (2013) argue that this feature is highly correlated with abstractness ratings (|*r*| = .51, *p* < 0.001; Hoffman et al. 2013), suggesting that abstract words, such as *set* or *time*, are more ambiguous than concrete words, such as *desk* or *orange*. Given the high correlation of ambiguity and abstractness, it is not surprising that svPPA patients produced more nouns that were abstract and ambiguous. It is also possible that svPPA patients produce nouns such as *furniture, object*, or *thing* that are superordinate in a hierarchically organized semantic network because they do not have access to more concrete words. These possibilities need to be studied in future work.

Previous work describing the hub-and-spoke model (Patterson, Nestor, & Rogers, 2007) claims that disease in the anterior temporal lobe is responsible for a universal semantic deficit in svPPA. We found in the present study that svPPA patients used verbs more frequently than patients with naPPA. A frequent use of a specific POS category does not necessarily reflect the integrity of the meaning of this word class. However, on the assumption that patients use words with which they are more familiar in a semistructured speech sample, the more frequent use of verbs than nouns in svPPA would be contrary to the claim that the meaning of all words is degraded in svPPA. Likewise, we have showed that the meaning of words for abstract nouns is relatively preserved in svPPA (Bonner et al., 2016; Cousins et al., 2016) and that the meaning of words dependent on number knowledge is relatively preserved in svPPA (Ash et al., 2016). In a longitudinal study of lexical expression in svPPA, we found progressively reduced use of concrete words relative to abstract words (Cousins et al., 2018). Findings such as these are more consistent with a relatively selective degradation of the lexicon in svPPA. Additional work is needed to assess these claims.

### 5.2 Lexical characteristics in naPPA

A distinguishing feature of naPPA is that these patients produced more speech errors and partial words than other groups did. The increased speech error and partial word rate in naPPA conforms to previous findings that naPPA patients exhibit effortful and non-fluent speech (Ash et al., 2013, 2009; Croot, Ballard, Leyton, & Hodges, 2012; Gorno-Tempini et al., 2004; Grossman et al., 1996; Weintraub et al., 1990). We related increased partial words and speech errors to cortical thinning in the left middle frontal gyrus, which is in line with previous findings (Ash et al., 2009; Gorno-Tempini et al., 2004; Grossman et al., 1996). An important characteristic of naPPA patients is their AoS, that is, the poor coordination of the motor articulators during speech production (Ash et al., 2009; Gorno-Tempini et al., 2011; Grossman et al., 1996, 2005; Josephs et al., 2006; Ogar et al., 2007). It is claimed that a subset of naPPA patients has AoS without grammatical impairments, and that this differs from naPPA patients with grammatical impairments who have AoS (Josephs et al., 2013, 2012). A major challenge to this area of investigation is the ability to detect speech errors in an objective, reliable and reproducible manner. A rating scale based on subjective judgments has been developed, but reliability is challenging (Josephs et al., 2012; Strand, Duffy, Clark, & Josephs, 2014). Another challenge is that partial words in naPPA patients are not explained solely by AoS. Additional work is needed to confirm the identification of speech errors and partial words in an naPPA cohort, to extend this observation to patients with movement disorders such as progressive supranuclear palsy and corticobasal syndrome, and to distinguish this from speech errors in patients with bulbar disease such as amyotrophic lateral sclerosis.

Patients with naPPA in our study produced fewer verbs than the other groups. Decreased verb use in naPPA patients has also been observed in previous studies (Ash et al., 2013, 2009). Several accounts have been forwarded to explain this finding. One suggestion is that naPPA patients have difficulty producing tense-inflected verbs and constructing complex sentence structures due to a syntactic deficit, which leads to a reduced use of verbs in their speech (Ash et al., 2013, 2009; Grossman et al., 1996; Grossman, Rhee, & Moore, 2005). Alternatively, disease in naPPA may also affect motor association regions of the frontal lobe and interfere with the representation of action knowledge associated with verbs of action (Hillis et al., 2004, 2002). Yet another possibility is that the entire class of verbs is associated with a richer and more demanding set of features—including not only its semantic attributes but also a rich set of grammatical and thematic properties—and naPPA patients have limitations in executive functioning that may make verbs more difficult for naPPA patients to process (Kramer et al., 2003; Libon et al., 2007; Weintraub, Rubin, & Mesulam, 1990). Previous work based on a smaller cohort of patients has suggested that the latter explanations are less likely than the grammatical one (Gunawardena et al., 2010), and we could not provide further evidence on these competing claims since the verb counts were not associated with cortical thinning in naPPA patients in our results. Additional work is needed to assess these claims.

### 5.3 Lexical characteristics in bvFTD

We hypothesized that bvFTD patients would differ in the counts of adjectives due to apathy and also that their nouns would be less abstract than those in the other groups. Our results showed that bvFTD patients produced fewer adjectives compared to controls, and their decreased adjective production was significantly correlated with their apathy scores, suggesting that bvFTD patients with fewer adjectives tended to be more apathetic. We identified regions of cortical thinning that were significantly related to apathy, including the left dorsolateral frontal and orbitofrontal gyri, and this result is in line with previous studies (Massimo et al., 2009, 2015). Furthermore, those regions that showed significant relations to the apathy scores were also significantly related to the adjective counts in bvFTD in our study. However, adverb production was not related to the degree of apathy in bvFTD. Also, we did not confirm our previous observation that bvFTD patients tend to produce relatively more concrete words than abstract words (Cousins et al., 2017), and this may have been due in part to the limited range of concreteness that could be achieved in a picture displaying many concrete nouns with little evocation of features leading to a description of the picture’s abstract characteristics.

It is interesting that adjective counts were negatively correlated with apathy scores in bvFTD, but adverb counts were not. This might be because not all adverbs serve as modifiers in a sentence. For example, so-called pro-adverbs, such as *here* or *there*, perform like function words, replacing prepositional phrases (e.g., *in the kitchen*). It might be the case that bvFTD patients used more pro-adverbs than modifying adverbs, resulting in an insignificant correlation with the apathy score. Additional work is needed to investigate this possibility.

Apathy is not only the most common symptom in bvFTD, occurring in 84% of patients (Rascovsky et al., 2011), but also a prevalent behavioral symptom in patients with other neurodegenerative disorders (e.g., Clark et al., 2008). Our study provided an easily reproducible language variable, adjective production, that might signal the degree of apathy in bvFTD patients. Identifying a language variable that is associated with apathy is particularly valuable, since social/behavioral impairments due to apathy cause the greatest caregiver distress (Massimo et al., 2009). Further study is needed to examine if adjective production is also associated with apathy in patients with other neurodegenerative diseases, such as Alzheimer’s disease.

### 5.4 Validating an automated lexical analysis of PPA patients’ speech

An important strength of our study is that we were able to validate an automated method for analyzing POS categories in a semi-structured speech sample produced by patients with speech deficits. An automated analysis is reliable in normal, healthy speakers. Here we were able to show that there was over 90% agreement between the automated POS tagging with the Google Universal scheme and the gold-standard POS tagging data of a linguist for speakers with abnormal speech. Indeed, the results of the present study are in line with many previous findings, suggesting that our novel, automated POS tagging and lexical analyses are valid in studying FTD patients’ speech.

Speech is central to human daily functioning and our approach has potential to serve as a clinical endpoint for treatment trials. While the present study focuses on cross-sectional data, work in progress assesses objective analyses of our longitudinal speech samples. Language production is a multifaceted process that requires a large expanse of brain tissue and is a sensitive marker for capturing even very early stages of neurodegeneration. Semi-structured speech data such as a picture description is inexpensive to collect on a large scale, when compared to MRI or lumbar puncture for cerebrospinal fluid which are expensive and/or invasive. However, it is nearly impossible to utilize and analyze large-scale speech data in a reproducible manner without an automated method. We believe that the method proposed in this paper can facilitate analyzing large-scale speech data in a quantifiable, automated, and reproducible way and can be used in automatic prescreening for neurodegeneration in the future (e.g., Cho et al. 2020).

## 6. Conclusion

While our study has many strengths, there are also some limitations that should be kept in mind when interpreting our results. One limitation is that the accuracy of the POS tagging for naPPA patients was not as high as for the other groups. Thus, the results of naPPA patients need to be interpreted with caution. This is an expected result for a POS tagger, since all existing POS taggers are trained with speech/text data of healthy adults. Accuracy could be improved if we trained a POS tagger using our patients’ speech samples with speech errors and other abnormalities as a training dataset. Also, since our automated methods rely on texts, there might be, for example, minor speech errors that were transcribed with regular spellings and our pipeline might have missed tagging those tokens as speech errors. We used an open-source POS tagger in the present study, but we plan to develop NLP tools, including a POS tagger, a syntactic dependency parser, and an automated speech recognition system for automatic transcription that will be trained on patients’ speech in the near future. Another limitation is that we had a relatively small number of digitized speech samples and a small number of MRI samples for naPPA patients. This limited our ability to perform statistically robust regression analyses in this patient group. We collect data on a regular basis, and future studies will contain more speech samples.

## Data Availability

Audio files are not sharable as voice is patient-identifiable information

## Appendices

**Table A:**
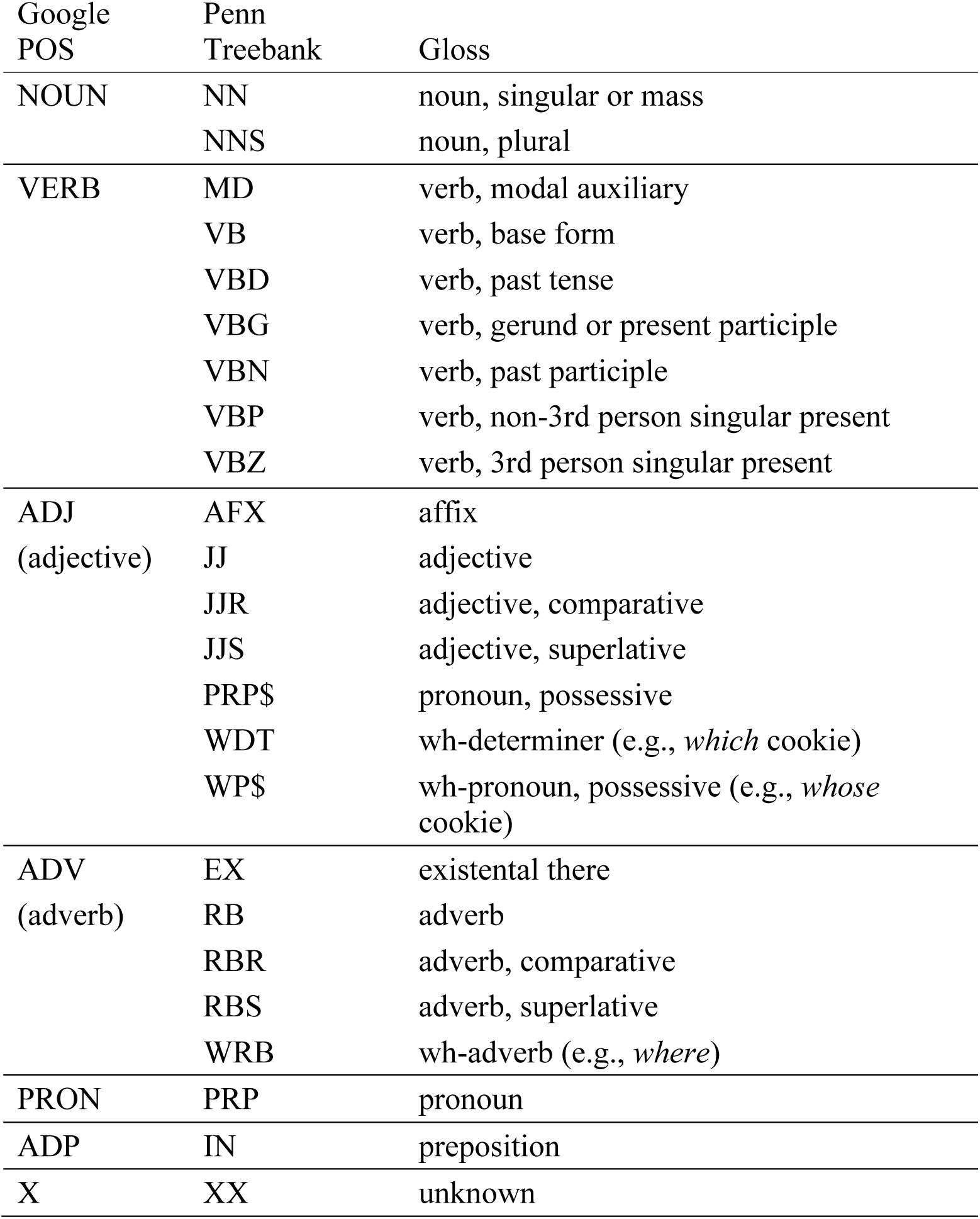
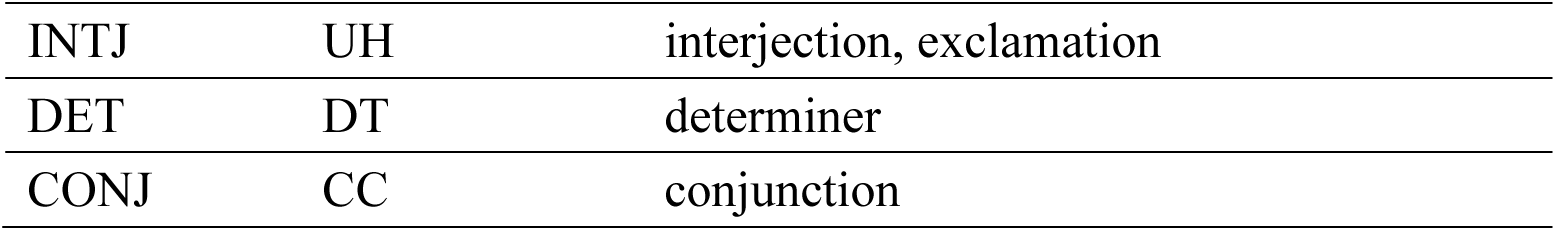
List of POS categories and mapping between the Google POS tag set and the Penn Treebank tag set. MD, VBD, VBP, and VBZ in the Penn Treebank tags were used to calculate the number of tense-inflected verbs.

| Google<br>POS | Penn<br>Treebank | Gloss |
| --- | --- | --- |
| NOUN | NN | noun, singular or mass |
|  | NNS | noun, plural |
| VERB | MD | verb, modal auxiliary |
|  | VB | verb, base form |
|  | VBD | verb, past tense |
|  | VBG | verb, gerund or present participle |
|  | VBN | verb, past participle |
|  | VBP | verb, non-3rd person singular present |
|  | VBZ | verb, 3rd person singular present |
| ADJ<br>(adjective) | AFX | affix |
|  | JJ | adjective |
|  | JJR | adjective, comparative |
|  | JJS | adjective, superlative |
| | PRP\$ | pronoun, possessive |
|  | WDT | wh-determiner (e.g., *which* cookie) |
| | WP\$ | wh-pronoun, possessive (e.g., *whose* cookie) |
| ADV<br>(adverb) | EX | existential there |
|  | RB | adverb |
|  | RBR | adverb, comparative |
|  | RBS | adverb, superlative |
|  | WRB | wh-adverb (e.g., *where*) |
| PRON | PRP | pronoun |
| ADP | IN | preposition |
| X | XX | unknown |
| INTJ | UH | interjection, exclamation |
| DET | DT | determiner |
| CONJ | CC | conjunction |

**Table B:** Demographic and clinical characteristics of the subset of patients with MRI data. The *p-*values for the group differences in this subset were from ANOVA analyses, except the sex ratio, where a chi-squared test was used. Student’s t-tests (all measures but the sex ratio) and a chi-squared test (sex ratio) were used for the comparisons of this subset with the full dataset. MMSE: Mini Mental State Exam; BNT: Boston Naming Test; PPT: Pyramids and Palm Trees Test; F: females; M: males.

|  | Controls<br>(n=18) | bvFTD<br>(n=42) | naPPA<br>(n=8) | svPPA<br>(n=26) | Group differences<br>in this subset | Comparison<br>with the full<br>set |
| --- | --- | --- | --- | --- | --- | --- |
| Age (years) | 65.9<br>(6.8) | 63 (8.5) | 65.5 (8.1) | 61.2<br>(7.1) | $F(3,90)=1.53$,<br>$p=0.21$ | $t(207)=-1.8$,<br>$p=0.07$ |
| Sex | 9 F, 9 M | 15 F,<br>27 M | 2 F, 6 M | 17 F, 9<br>M | $\chi^2=7.26$, $p=0.06$ | $\chi^2=0.12$,<br>$p=0.72$ |
| Education<br>(years) | 16.1<br>(2.9) | 15.9<br>(2.2) | 17.4 (3) | 15.3<br>(2.6) | $F(3,90)=1.37$,<br>$p=0.26$ | $t(207)=0.9$,<br>$p=0.38$ |
| Disease<br>duration<br>(years) | - | 4 (3.4) | 3 (2) | 3.6 (2) | $F(2,73)=0.43$,<br>$p=0.66$ | $t(158)=-0.7$,<br>$p=0.47$ |
| MMSE (0-30) | 28.9<br>(1.1) | 25.1<br>(4.3) | 25.1 (3.4) | 23.6<br>(6.1) | $F(3,88)=5.26$,<br>$p=0.002$ | $t(222)=1.9$,<br>$p=0.06$ |
| BNT (0-30) | 27.7<br>(2.7) | 24.5<br>(4.1) | 24.8 (5.1) | 7.7 (6.3) | $F(3,89)=90.81$,<br>$p<0.001$ | $t(200)=0.3$,<br>$p=0.76$ |
| PPT (0-52) | 51.3<br>(1.1) | 45.4<br>(6.9) | 48.5 (3.7) | 39.1<br>(7.1) | $F(3,48)=8.75$,<br>$p<0.001$ | $t(111)=0.4$,<br>$p=0.68$ |
| Animals and<br>tools (max 60<br>secs) | 16.8 (5) | 10 (4.9) | 9.8 (4.8) | 6 (3.9) | $F(3,86)=18.52$,<br>$p<0.001$ | $t(190)=1.3$,<br>$p=0.2$ |

**Table C:**
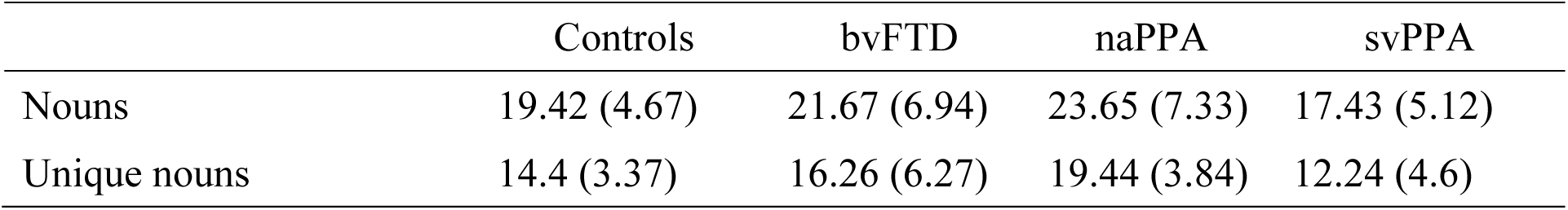
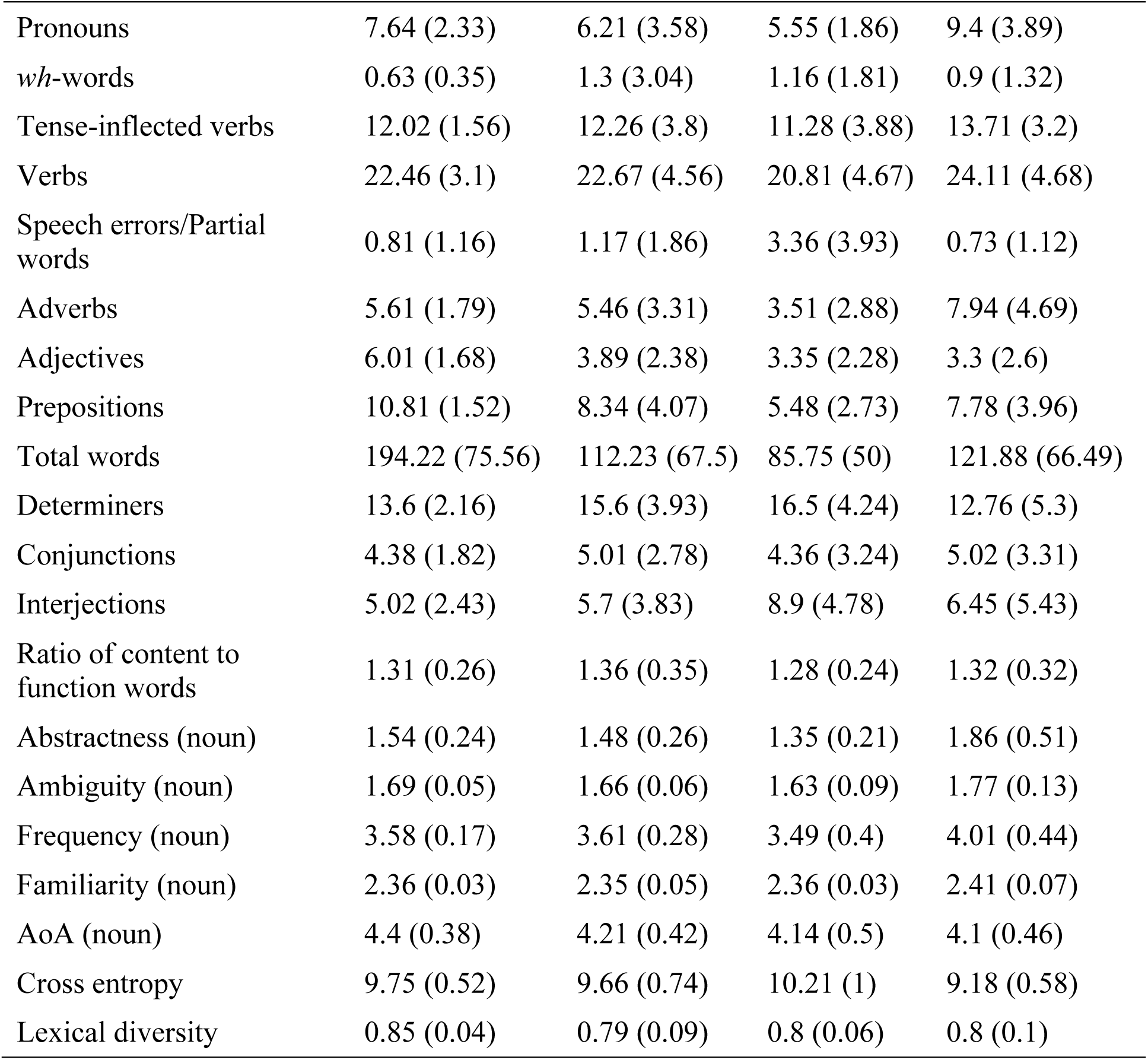
POS counts per 100 words and lexical measures of the subset of patients with MRI data.

|  | Controls | bvFTD | naPPA | svPPA |
| --- | --- | --- | --- | --- |
| Nouns | 19.42 (4.67) | 21.67 (6.94) | 23.65 (7.33) | 17.43 (5.12) |
| Unique nouns | 14.4 (3.37) | 16.26 (6.27) | 19.44 (3.84) | 12.24 (4.6) |
| Pronouns | 7.64 (2.33) | 6.21 (3.58) | 5.55 (1.86) | 9.4 (3.89) |
| *wh*-words | 0.63 (0.35) | 1.3 (3.04) | 1.16 (1.81) | 0.9 (1.32) |
| Tense-inflected verbs | 12.02 (1.56) | 12.26 (3.8) | 11.28 (3.88) | 13.71 (3.2) |
| Verbs | 22.46 (3.1) | 22.67 (4.56) | 20.81 (4.67) | 24.11 (4.68) |
| Speech errors/Partial words | 0.81 (1.16) | 1.17 (1.86) | 3.36 (3.93) | 0.73 (1.12) |
| Adverbs | 5.61 (1.79) | 5.46 (3.31) | 3.51 (2.88) | 7.94 (4.69) |
| Adjectives | 6.01 (1.68) | 3.89 (2.38) | 3.35 (2.28) | 3.3 (2.6) |
| Prepositions | 10.81 (1.52) | 8.34 (4.07) | 5.48 (2.73) | 7.78 (3.96) |
| Total words | 194.22 (75.56) | 112.23 (67.5) | 85.75 (50) | 121.88 (66.49) |
| Determiners | 13.6 (2.16) | 15.6 (3.93) | 16.5 (4.24) | 12.76 (5.3) |
| Conjunctions | 4.38 (1.82) | 5.01 (2.78) | 4.36 (3.24) | 5.02 (3.31) |
| Interjections | 5.02 (2.43) | 5.7 (3.83) | 8.9 (4.78) | 6.45 (5.43) |
| Ratio of content to function words | 1.31 (0.26) | 1.36 (0.35) | 1.28 (0.24) | 1.32 (0.32) |
| Abstractness (noun) | 1.54 (0.24) | 1.48 (0.26) | 1.35 (0.21) | 1.86 (0.51) |
| Ambiguity (noun) | 1.69 (0.05) | 1.66 (0.06) | 1.63 (0.09) | 1.77 (0.13) |
| Frequency (noun) | 3.58 (0.17) | 3.61 (0.28) | 3.49 (0.4) | 4.01 (0.44) |
| Familiarity (noun) | 2.36 (0.03) | 2.35 (0.05) | 2.36 (0.03) | 2.41 (0.07) |
| AoA (noun) | 4.4 (0.38) | 4.21 (0.42) | 4.14 (0.5) | 4.1 (0.46) |
| Cross entropy | 9.75 (0.52) | 9.66 (0.74) | 10.21 (1) | 9.18 (0.58) |
| Lexical diversity | 0.85 (0.04) | 0.79 (0.09) | 0.8 (0.06) | 0.8 (0.1) |

